# Extracardiac Thoracoabdominal Atherosclerosis in Heart Transplant Candidates is not Associated with Standard Modifiable Cardiovascular Risk Factors

**DOI:** 10.64898/2026.02.25.26347056

**Authors:** Thomas Readford, Martin Ugander, Peter L. Kench, Christopher Hayward, Gemma Figtree, James Nadel, Nicola Giannotti

## Abstract

**Background:** Screening for atherosclerosis focuses on identifying Standard Modifiable Risk Factors (SMuRFs), including diabetes, hypertension, hyperlipidaemia, and smoking.

**Purpose:** To compare the extracardiac thoracoabdominal atherosclerotic plaque burden, as measured by computed tomography angiography (CTA), among heart transplant candidates with ischemic or non-ischemic cardiomyopathy (ICM, NICM), and evaluate potential associations between plaque burden and SMuRFs.

**Methods:** This retrospective study identified heart transplant candidates with ICM or NICM matched for age and sex, undergoing thoracoabdominal CTA. Patients were classified as with SMuRFs or SMuRF-less. Extracardiac thoracoabdominal non-calcified and calcified atherosclerotic plaque was classified as present or absent across 78 arterial segments per patient.

**Results:** Among included patients (n=167, median [interquartile range] age 58 [53-63] years, 16% female, 51% NICM), 40 patients (24%) were SMuRF-less (ICM: 16/82 (20%), NICM: 24/85 (28%), age 56 [50-67] years). Overall, out of 13,026 arterial segments analysed, 1,746 (13%) were affected by atherosclerotic plaque (9 [4-15] segments per patient). ICM had a higher total plaque burden than NICM (11 [7-18] vs 6 [3-11] segments per patient, p<0.001). SMuRF-less patients showed no difference in non-calcified, calcified, or total plaque burden compared to patients with SMuRFs, among all patients (ICM+NICM, p>0.17) and within the ICM and NICM groups, respectively (p>0.30).

**Conclusions:** The burden of extracardiac thoracoabdominal atherosclerotic plaque is higher among heart transplant candidates with ICM. However, it does not differ between SMuRF-less or those with SMuRFs, regardless of underlying ICM or NICM. The prevalence of SMuRFs is not an effective marker to determine the need to screen for extracardiac atherosclerotic plaque among heart transplant candidates.

**GRAPHICAL ABSTRACT:** 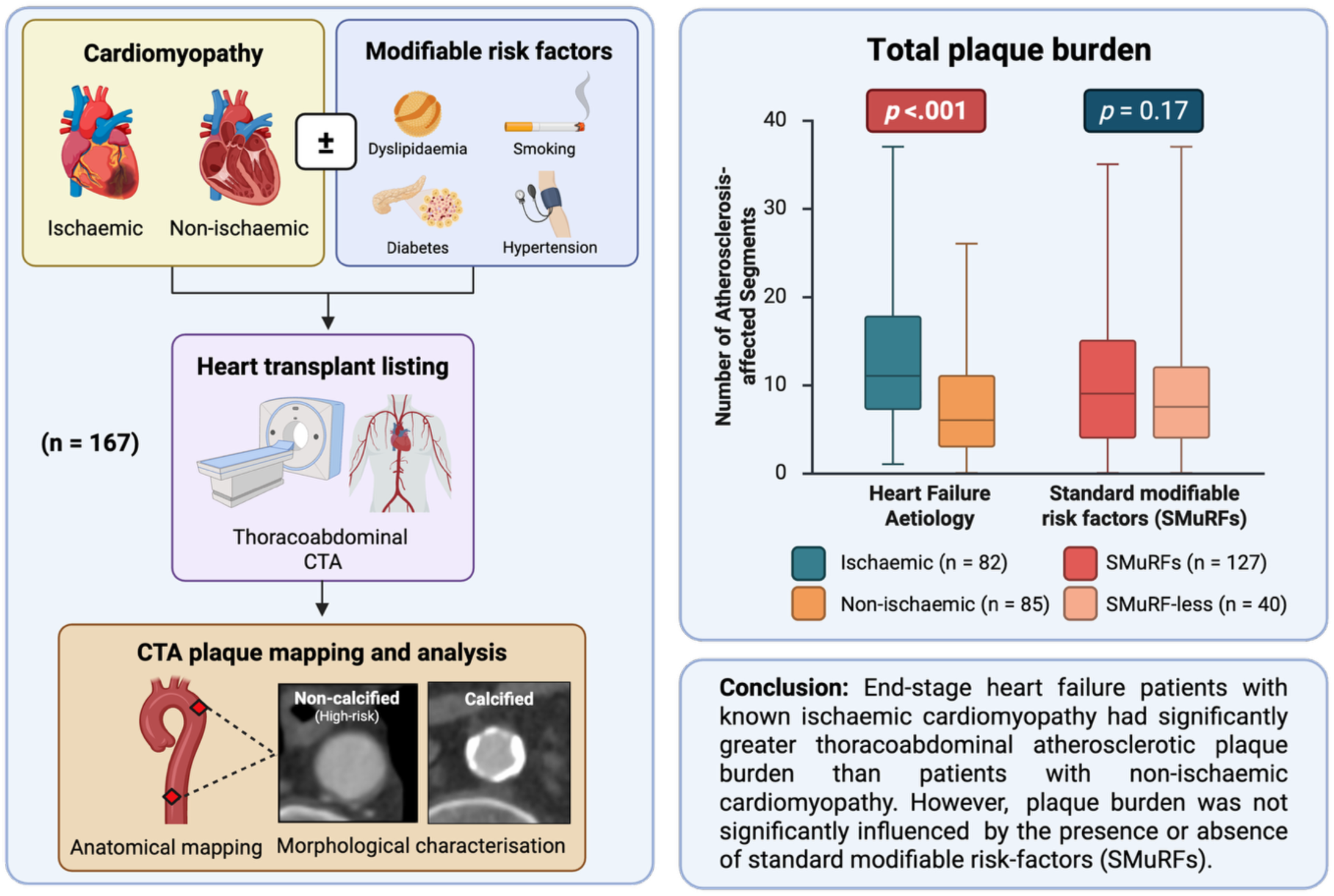

## INTRODUCTION

Atherosclerotic cardiovascular disease (ASCVD) remains the leading cause of morbidity and mortality globally. (1) A significant proportion of patients who experience major adverse cardiovascular and cerebrovascular events (MACCE) have very few or zero standard modifiable risk factors (SMuRFs), including hypertension, diabetes mellitus, dyslipidaemia, and smoking. (2, 3) This phenotype has been termed “SMuRF-less.” (2, 3) Hence, the occurrence of MACCE due to atherosclerosis in these apparently low-risk individuals highlights important limitations of current risk factor-based prediction models and potential screening strategies. (2, 3) This growing evidence of a disconnect between traditional SMuRFs and clinical outcomes has renewed interest in identifying alternative drivers of atherosclerosis disease progression that may operate independently of conventional modifiable risk factors.

Beyond traditional risk factor assessment, direct imaging of atherosclerosis-prone territories such as the coronary and carotid arteries can identify high-risk plaque features that predispose atherosclerotic lesions to rupture and substantially improve the prediction of MACCE, as demonstrated in large-scale prospective trials including the SCOT-HEART and the Rotterdam studies. (4, 5) Therefore, given the systemic nature of atherosclerosis, the burden of extracardiac atherosclerosis across many other vascular beds may similarly serve as a potentially useful indicator of future risk. (6, 7) In this context, thoracoabdominal computed tomography angiography (CTA) offers a relatively simple, widely available, and non-invasive method to evaluate extracardiac atherosclerotic disease throughout the aorta and its major thoracoabdominal branches. (6) Therefore, the presence and extent of potentially high-risk extracardiac atherosclerotic plaque detected by CTA should be interrogated, when possible, as it may represent a valuable imaging biomarker of clinically significant disease, even in patients that are SMuRF-less.

In patients with end-stage cardiomyopathy from ischaemic or non-ischaemic causes evaluated for heart transplantation, thoracoabdominal CTA is routinely performed to exclude malignancy before listing and immunosuppression. Hence, this diagnostic workup may provide an opportunity to assess extracardiac atherosclerotic plaque burden, a marker of atherosclerotic disease progression in the coronary and carotid arterial beds, without additional imaging, thereby guiding medical optimisation pre- and post-transplant. (7) Therefore, this study aimed to: (i) compare the extent of extracardiac thoracoabdominal atherosclerotic plaque on pre-transplant CTA between patients with ischaemic cardiomyopathy (ICM) and those with non-ischaemic cardiomyopathy (NICM), and (ii) examine whether extracardiac thoracoabdominal atherosclerotic plaque burden is associated with traditional modifiable cardiovascular risk factors.

## METHODS

### Study design and ethics

This study was a single-centre retrospective observational cohort study approved by the local human subject research ethics committee with a retrospective waiver of individual informed consent (REF: SVH/ETH02366).

### Clinical site and population

The study was conducted at a tertiary referral centre in metropolitan Sydney, New South Wales, Australia. As one of the few accredited heart transplant services in the country, the site receives referrals for advanced heart failure management and transplantation from bordering state health services, including the Australian Capital Territory, the Northern Territory, South Australia and Tasmania.

### Participant recruitment

Participants were retrospectively recruited from an existing database of orthotopic heart transplant recipients.

### Inclusion criteria

Participants were included if they underwent CTA of the entire aorta and its thoracic and abdominal branches to the level of the aortoiliac bifurcation. Participants were excluded if no contrast-enhanced CTA imaging was available before transplant. Inclusion was limited to patients who underwent heart transplant between 2012 and 2025 due to the availability of electronic databases of both imaging and digital medical records. Patients with peripartum cardiomyopathy with a concurrent history of spontaneous coronary artery dissection were excluded from inclusion in this study, as they could not easily be allocated to the ICM or NICM groups. Patients who had previously undergone orthotopic heart transplant were also excluded.

### Data collection

Clinical and CTA imaging data were collected over five months from May to September 2025. Study data were collected and managed using a customised REDCap electronic data capture tool hosted at The University of Sydney. (8, 9) Clinical notes, discharge summaries and other correspondence were reviewed to capture relevant medication, clinical history and cardiovascular risk factor data. Baseline blood chemistry, lipid profiles, and diabetes biomarkers were recorded at time points that did not align with acute heart failure presentations. Data were then exported from REDCap to Microsoft Excel (Microsoft Corporation, Redmond, WA, USA) for data cleaning and preparation prior to subsequent statistical analyses. Anonymised clinical images were stored securely on a dedicated research database at the University of Sydney.

### Allocation

Determination of patients’ antecedent heart failure aetiology as ischaemic or non-ischaemic was made through a combination of medical record review, echocardiography and coronary angiography reports, where available. When clinical notes recorded ischaemic cardiomyopathy from coronary atherosclerosis with a history of regional wall motion abnormalities on transthoracic echocardiography, patients were classified into the ICM group. Where clinical notes recorded that patients’ heart failure was caused by non-ischaemic causes, or ischaemic heart disease caused by a non-atherosclerotic mechanism (i.e. iatrogenic coronary dissection), patients were classified into the NICM group. Patients with non-obstructive coronary artery disease (CAD) confirmed by angiography using the CAD-RADS classification system were allocated to the NICM group. After determining that patients were eligible for inclusion, patients in the NICM group were matched for age and sex with patients with ICM.

### Image acquisition

Thoracoabdominal CTA data from individuals undergoing the diagnostic workup for heart transplantation were retrospectively collected. Following a low-dose scanogram of the torso, patients received 75–100 mL of Ultravist 300 iodinated contrast agent (Bayer, Leverkusen, Germany), administered via a power injector at 3 mL/sec. After a 20-second delay, the CTA acquisition was triggered by bolus tracking technique using a circular region of interest at the level of the aortic arch set at 150 Hounsfield units (HU), or in case of patients with low ejection fraction, where the operator deemed that enhancement was best achievable. Helical CTA acquisition was performed extending from the base of the neck to below the iliac crest. Tube voltage was ranged between 100 to 120 kilovolts. Automatic tube current modulation was used in all cases. The scanning protocols are included in the supplementary material in the Appendix.

### Image analysis and multi-planar reconstruction

A semi-quantitative approach was used to map the plaque distribution in the thoracoabdominal area and quantify total plaque burden. Image analysis was conducted using Osirix MD V14.1.2 (Pixmeo, Geneva, Switzerland). Images were analysed by a single-reader (TR), who was trained in atherosclerotic plaque phenotype classification using CTA. A randomly selected sample of cases (n=25) were reviewed by a second reader (NG) with >10 years in plaque imaging to confirm consistency in methodology and accuracy in plaque characterisation. Overall image quality was confirmed prior to analysis of each CTA case using a Likert scale to confirm sufficient diagnostic quality of each CTA. Image quality was deemed good, fair, or poor based on the resolvability of vascular structures, arterial contrast enhancement, and the presence of significant metallic artefact from implanted devices.

Studies with poor image quality were subsequently excluded from analysis. Scans with a field of view which did not include the entire chest and abdomen to the level of the aortoiliac bifurcation were not included in the analysis. A consistent range of window width and level to improve contrast differentiation of vessel wall, lumen and plaque phenotype (calcific vs non calcific) was used to analyse all images (width: 200-250 HU, level: 800-1000 HU). Curved and orthogonal multi-planar reformatting was applied to all CTA data to facilitate vessel and plaque analysis across the arterial structures. The centreline was drawn manually through the longitudinal axis of the aorta and branch vessels as listed in table 1. Consequently, the reconstructed CTA data were visually assessed in the reconstructed axial plane to identify atherosclerosis-affected arterial segments.

**Table 1:**
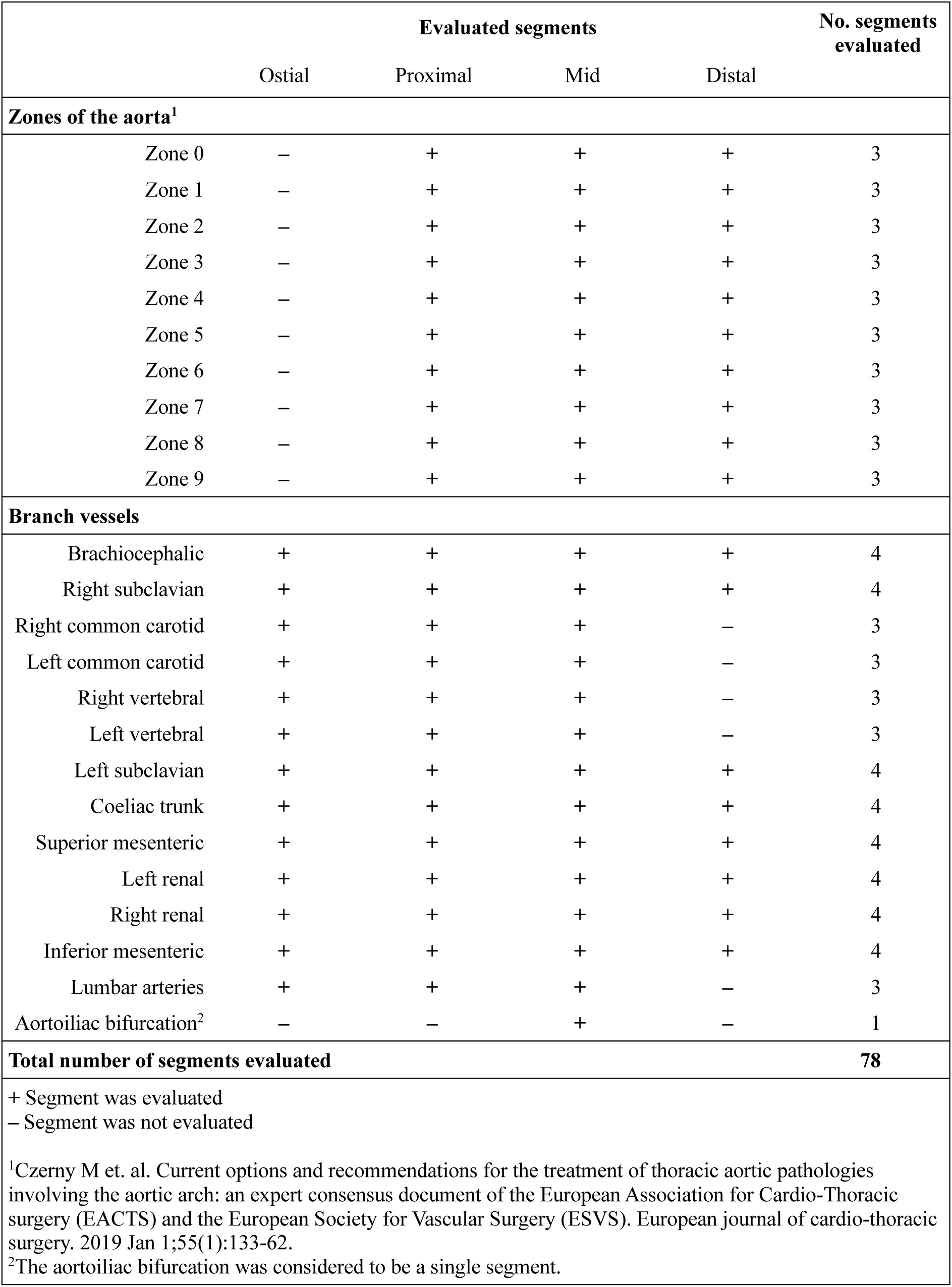
CTA-assessable aortic zones and branch arterial segments which were evaluated.

|  | Evaluated segments |  |  |  | No. segments evaluated |
| --- | --- | --- | --- | --- | --- |
|  | Ostial | Proximal | Mid | Distal |  |
| Zones of the aorta<sup>1</sup> |  |  |  |  |  |
| Zone 0 | – | + | + | + | 3 |
| Zone 1 | – | + | + | + | 3 |
| Zone 2 | – | + | + | + | 3 |
| Zone 3 | – | + | + | + | 3 |
| Zone 4 | – | + | + | + | 3 |
| Zone 5 | – | + | + | + | 3 |
| Zone 6 | – | + | + | + | 3 |
| Zone 7 | – | + | + | + | 3 |
| Zone 8 | – | + | + | + | 3 |
| Zone 9 | – | + | + | + | 3 |
| Branch vessels |  |  |  |  |  |
| Brachiocephalic | + | + | + | + | 4 |
| Right subclavian | + | + | + | + | 4 |
| Right common carotid | + | + | + | – | 3 |
| Left common carotid | + | + | + | – | 3 |
| Right vertebral | + | + | + | – | 3 |
| Left vertebral | + | + | + | – | 3 |
| Left subclavian | + | + | + | + | 4 |
| Coeliac trunk | + | + | + | + | 4 |
| Superior mesenteric | + | + | + | + | 4 |
| Left renal | + | + | + | + | 4 |
| Right renal | + | + | + | + | 4 |
| Inferior mesenteric | + | + | + | + | 4 |
| Lumbar arteries | + | + | + | – | 3 |
| Aortoiliac bifurcation<sup>2</sup> | – | – | + | – | 1 |
| Total number of segments evaluated |  |  |  |  | 78 |
| + Segment was evaluated |  |  |  |  |  |
| – Segment was not evaluated |  |  |  |  |  |

<sup>1</sup>Czerny M et. al. Current options and recommendations for the treatment of thoracic aortic pathologies involving the aortic arch: an expert consensus document of the European Association for Cardio-Thoracic surgery (EACTS) and the European Society for Vascular Surgery (ESVS). European journal of cardio-thoracic surgery. 2019 Jan 1;55(1):133-62.
<sup>2</sup>The aortoiliac bifurcation was considered to be a single segment.

### Plaque characterisation and anatomical mapping

In total, 78 arterial segments for each CTA case were evaluated for atherosclerotic disease distribution and morphology. **(Table 1)**

Aortic zones were defined using the Ishimaru classification system, as endorsed by the EACTS and ESVS 2019 expert consensus statement for pathologies of the aortic arch (10, 11). Each zone was subdivided into proximal, mid, and distal segments, as seen in **Figure 1**. Arterial branch vessels were divided into ostial, proximal, mid, and distal, with the aortoiliac bifurcation assessed as a single segment and common carotid arteries evaluated only to the mid-vessel because of CTA field-of-view limitations.

**Figure 1:**
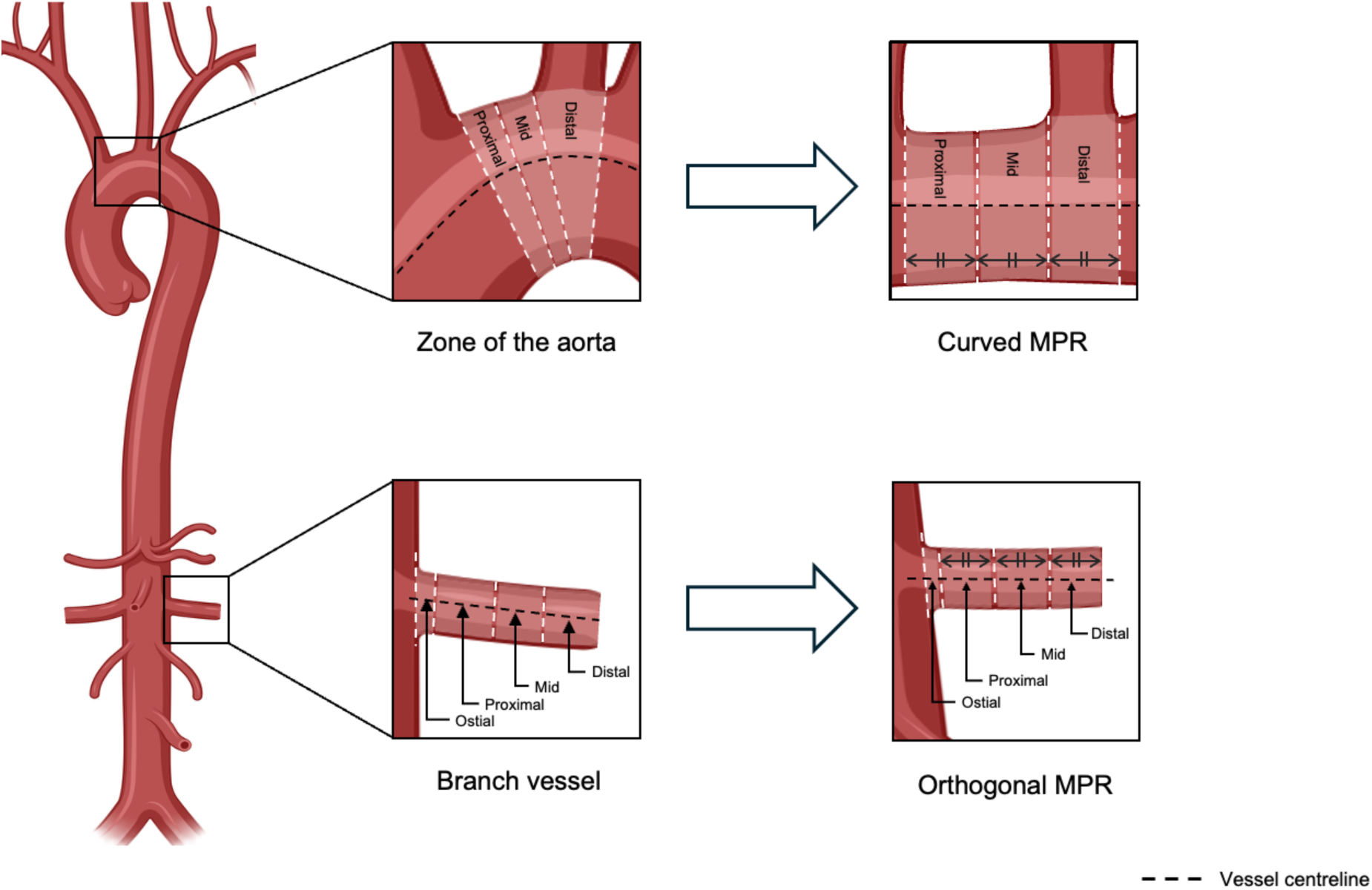
Overview of the method used to define and reconstruct arterial segments. Zones of the aorta were defined using the ‘Ishimaru’ classification system.

For each arterial segment analysed, atherosclerotic disease was determined to be present or not present. If plaque was present, it was morphologically characterised by the reader as soft (low attenuation), mixed (partially calcified) or calcific (entirely calcified). Plaques which were not entirely calcified i.e. soft, mixed or ruptured, were classified as non-calcified or high-risk plaque. **(Figure 2)**

**Figure 2:**
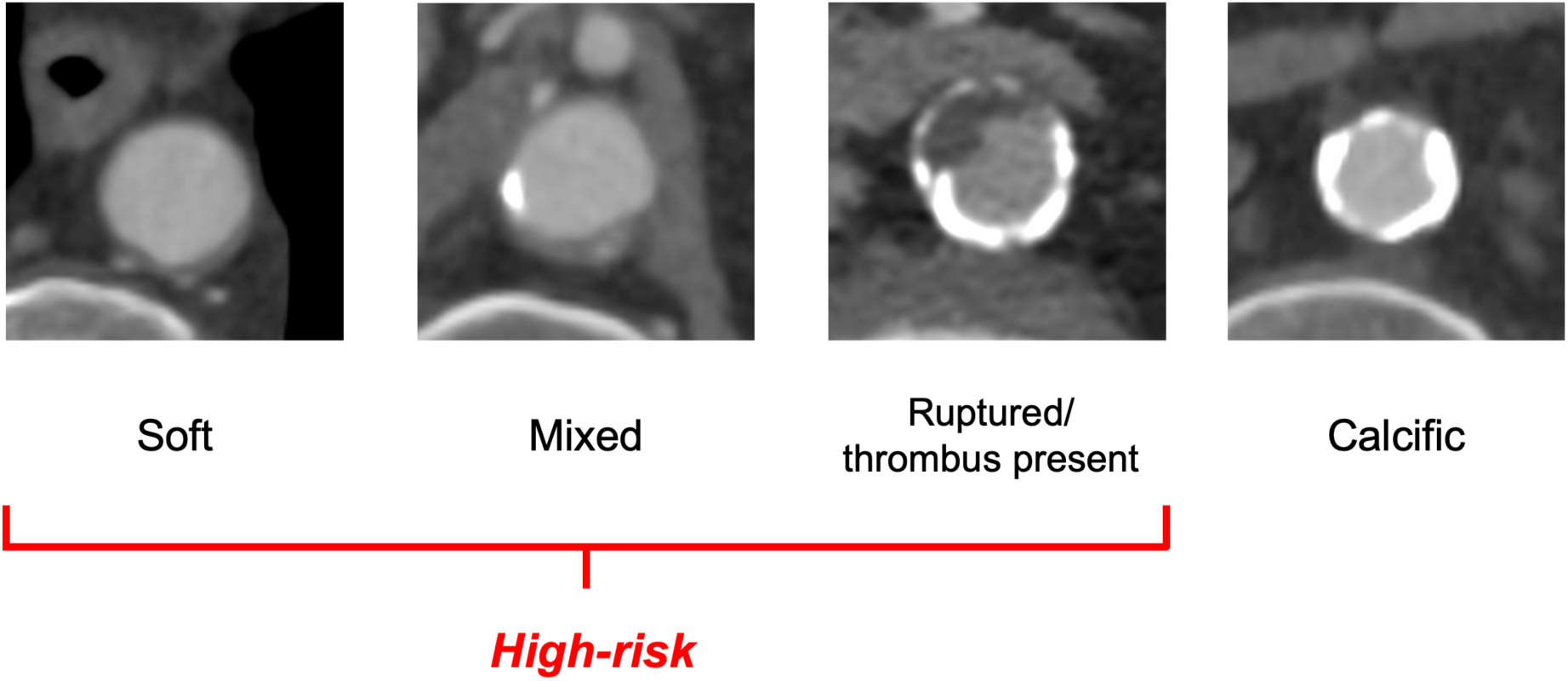
Typical CTA appearances of soft, mixed and calcific plaque.

Where the extent of the atherosclerosis extended beyond the boundaries of one segment and spilled over to an adjacent segment, both segments were considered to be affected by atherosclerosis. Total plaque burden was quantified as the number of visualised arterial segments affected by atherosclerosis. Non-calcified plaque burden was calculated as the number of arterial segments affected by soft or mixed plaque ?or the presence of high-risk plaque.

### Statistical analysis

Statistical analysis was completed using IBM SPSS Statistics V29.0.2.0 (IBM Corp., Armonk, NY, USA). Data are presented as mean±SD or median [interquartile range]. The Shapiro-Wilks test, along with a visual inspection of the data distribution, was used to determine whether the data were normally distributed. When data were normally distributed, the parametric independent-samples t-test was used to test for differences. When data were not normally distributed, the nonparametric Mann-Whitney U test and the Kruskal-Wallis test were used to test for differences. Multivariate linear regression analysis was performed to evaluate the potential effects of residual risk factors on plaque burden. Intergroup differences in plaque distribution and composition were assessed using Quade’s rank analysis of covariance (ANCOVA).

## RESULTS

### Of 576 screened patients, 167 were included in this study. **(Figure 3)**

Of the 167 patients included, 26 (16%) were female. A total of 82 (49%) patients had ICM, and 85 (51%) of patients had NICM. The median age at time of CTA was 58 [53-63] years. Within the NICM group, a total of 17 (20%) patients had NO-CAD. In patients with known obstructive coronary disease, the age at first occlusion, myocardial infarction, or revascularisation attempt was 51 [43-57] years and ranged from 22 years to 71 years. A total of 40 (24%) subjects from the initial 167 included patients were SMuRF-less before heart transplantation (ICM: 16/82 (20%), NICM 24/85 (28%), age 56 [50-67] years). **(Table 2)**

**Figure 3:**
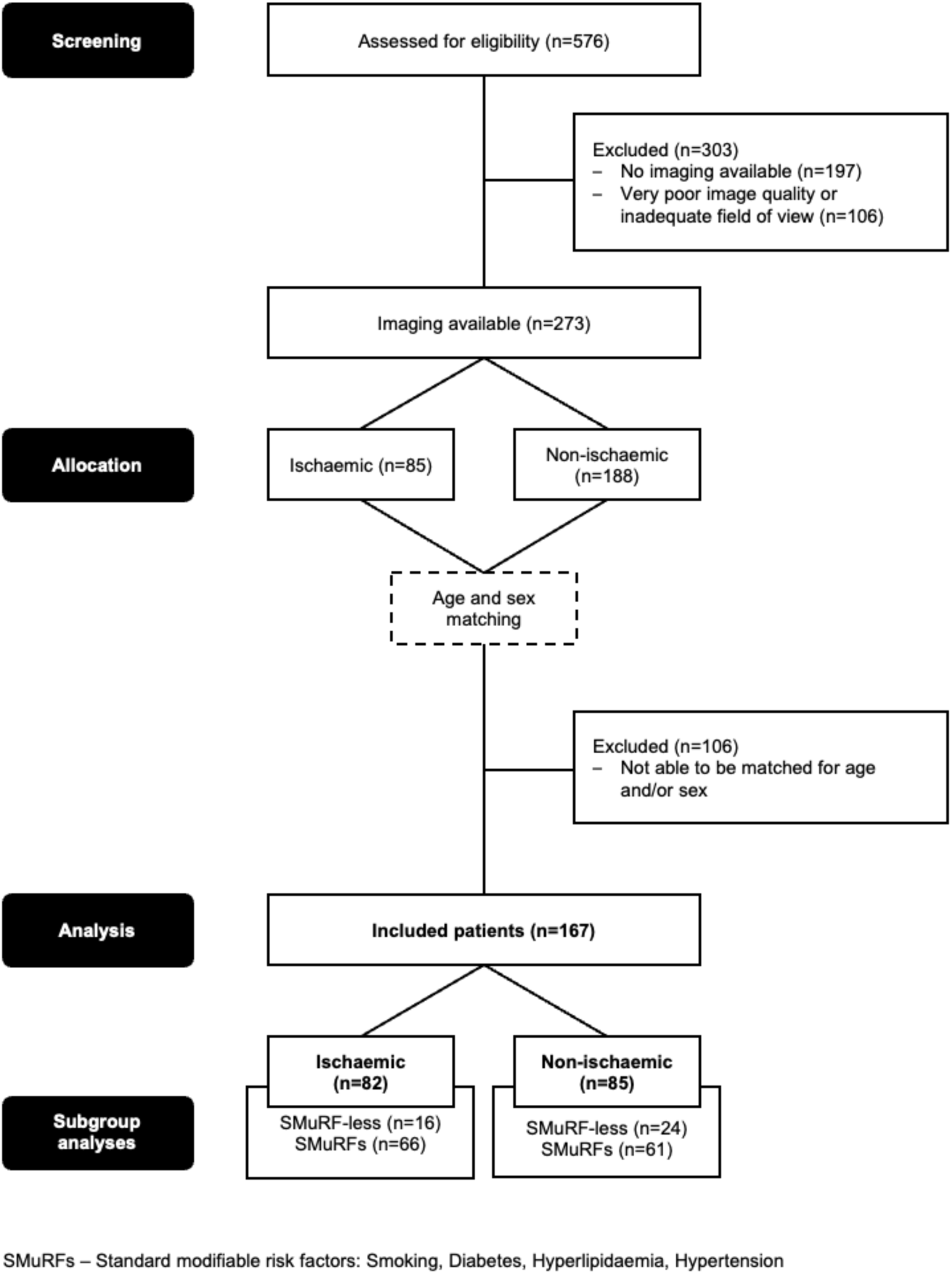
Flow chart of patient selection for inclusion in this study.

**Table 2:**
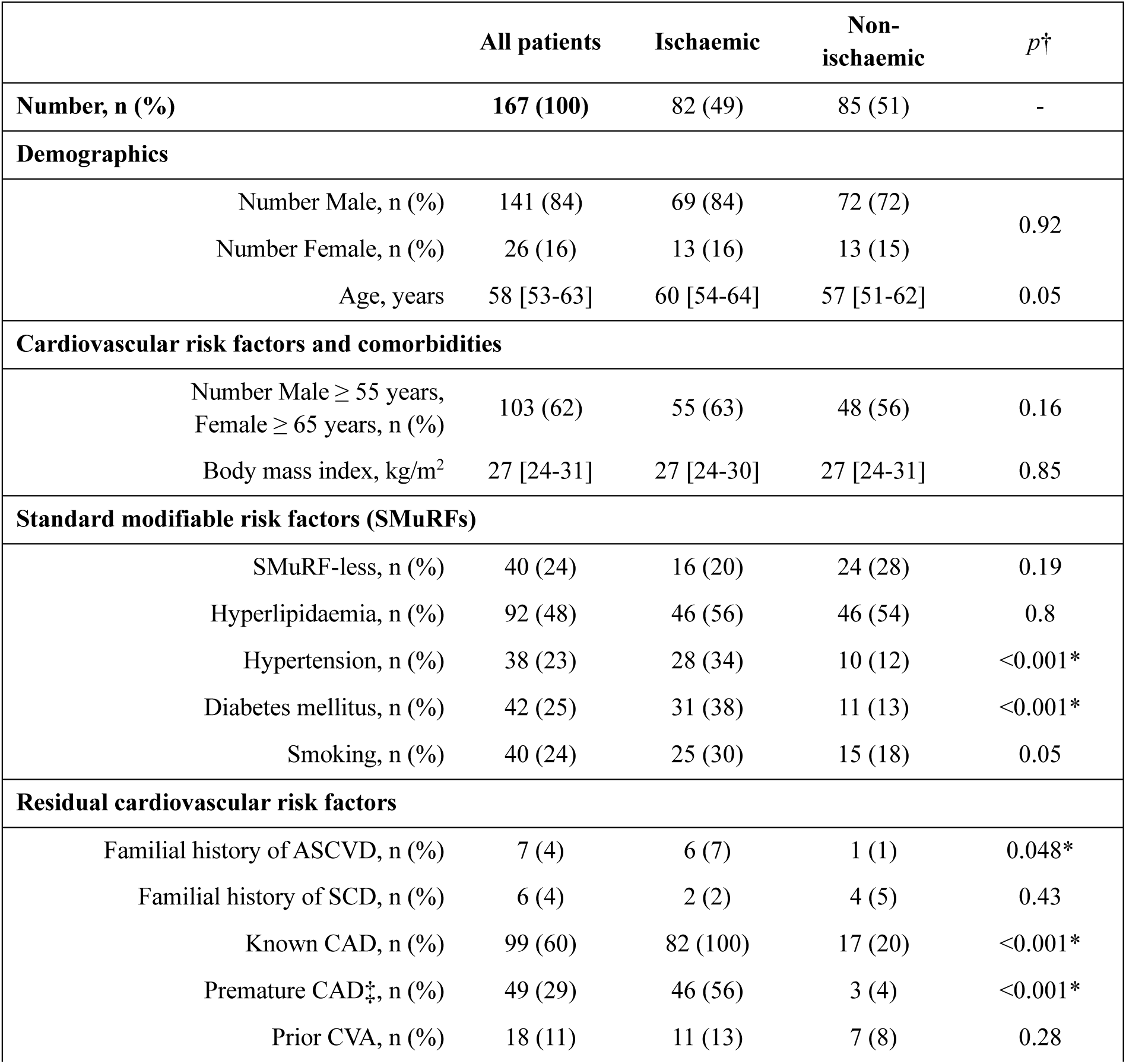
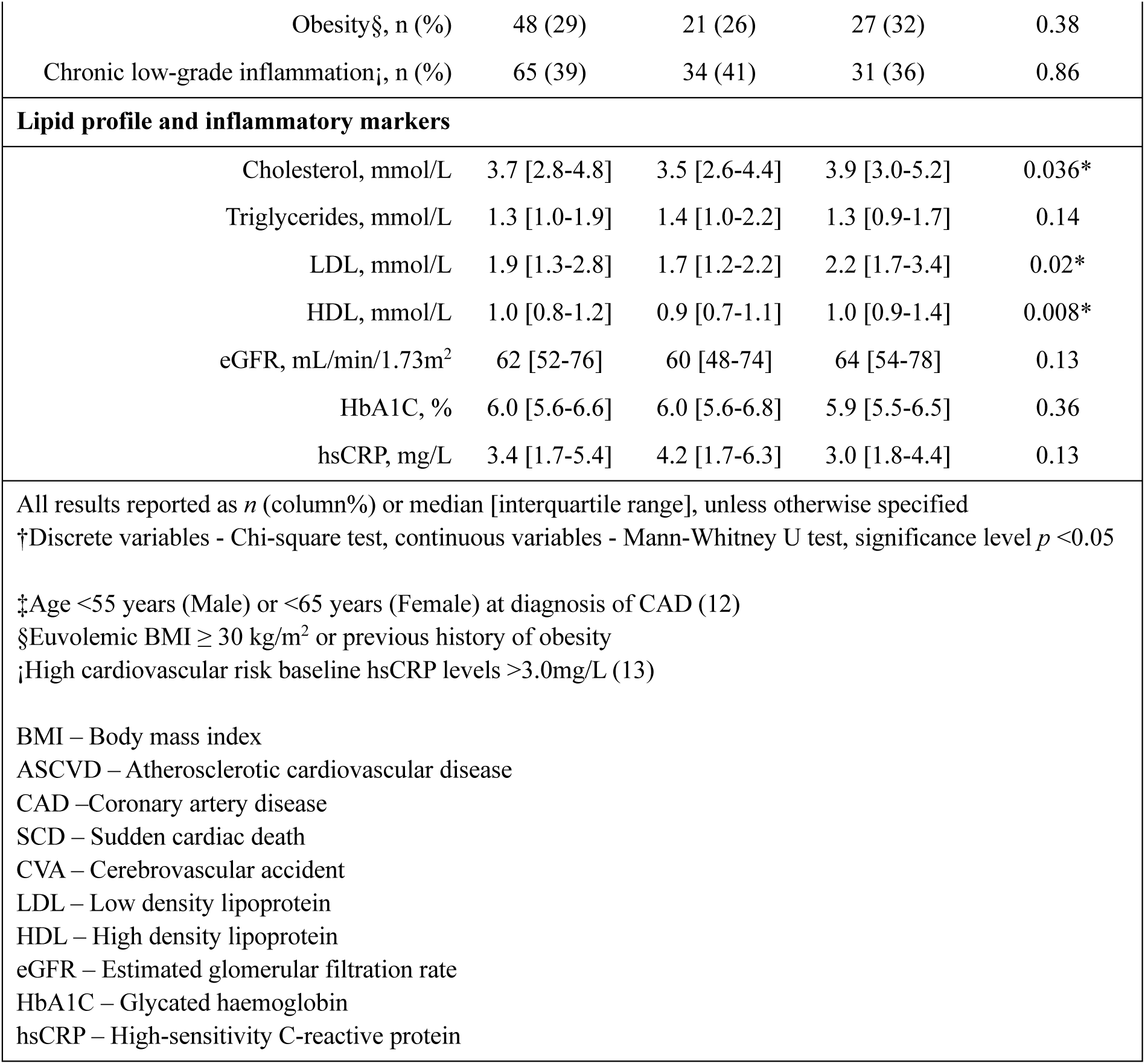
Patient demographics and comorbidities.

A total of 13,026 arterial segments were evaluated across 167 cases to determine whether atherosclerosis was present. Compared with patients with NICM, patients with ICM had a greater total plaque burden (number of atherosclerotic segments affected: 11 [7-18] vs 6 [3-11], p<0.001). **(Figure 4)** Furthermore, patients with ICM had a greater number of arterial segments affected by soft, mixed and calcific plaque types (*p*<0.05 for all) **(Table 4) (Figure 5).** There was no difference in total, non-calcified or calcified plaque burden between male and female patients (*p*>0.05 for all).

**Figure 4:**
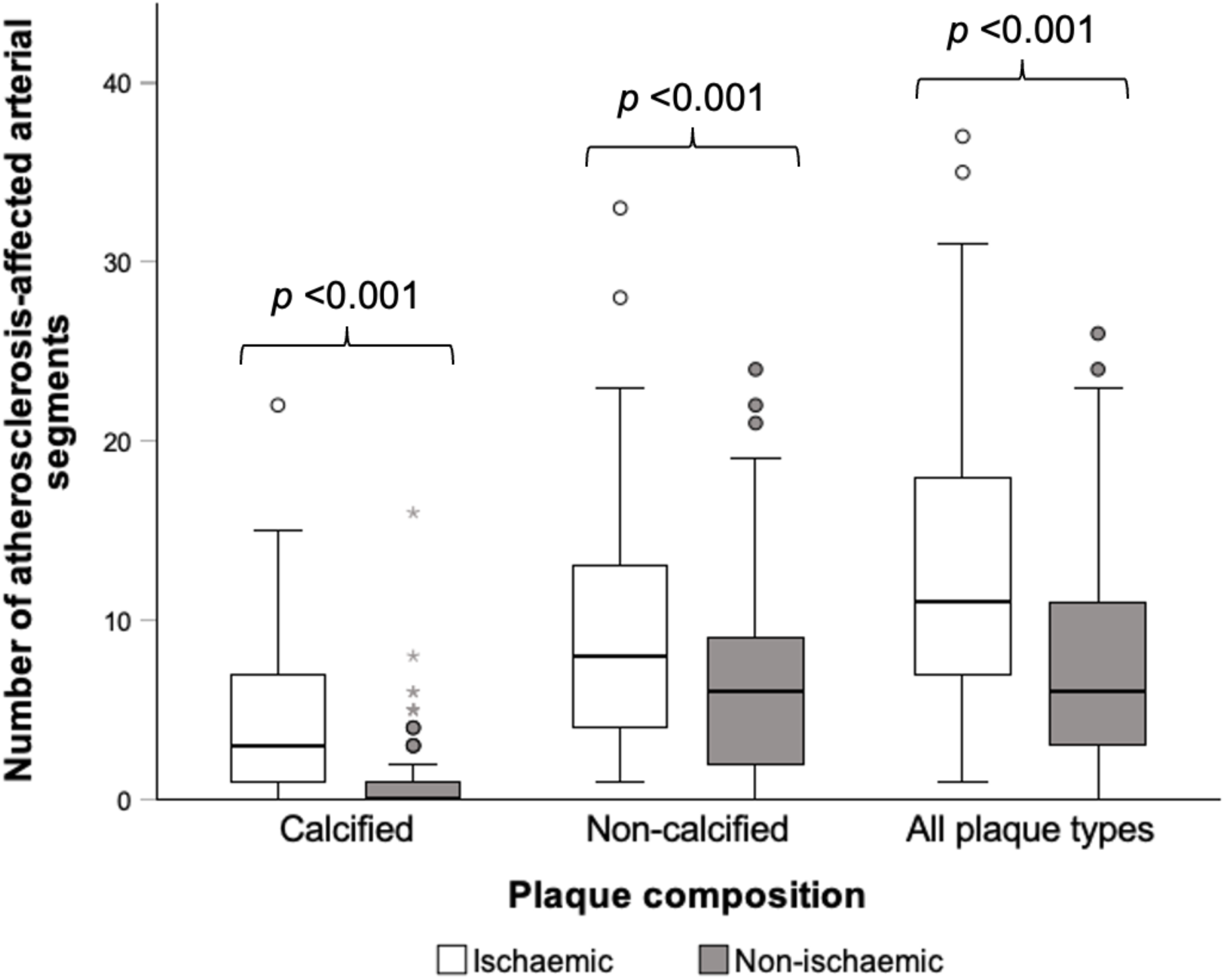
Non-calcified, calcified and total plaque burden in patients with ischaemic CM vs. non-ischaemic CM.

**Figure 5:**
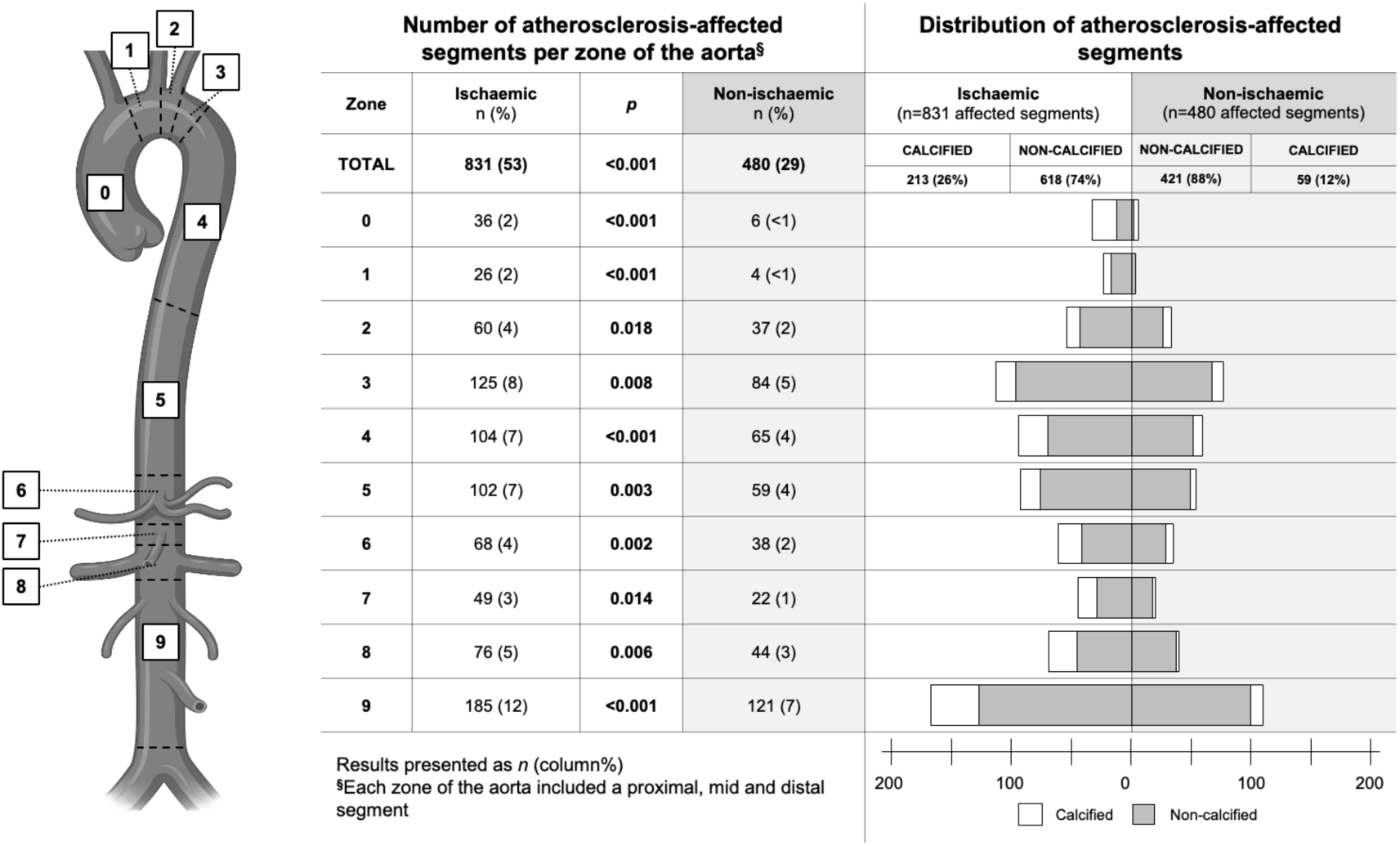
Distribution of atherosclerosis-affected arterial segments in the aorta, ICM vs. NICM groups.

**Table 4:** Number of atherosclerosis-affected segments, ICM vs. NICM groups.

|  | All patients<br>(n = 167) | Ischaemic<br>(n = 82) | Non-ischaemic<br>(n = 85) | *p* |
| --- | --- | --- | --- | --- |
| Total number of segments evaluated, n (%) | 13,026 (100) | 6,396 (49) | 6,630 (51) | - |
| Number of affected segments, n (%) | 1,826 (14) | 1,106 (17) | 720 (11) | <0.001* |
| Number of affected segments, per patient | 9 [4-15] | 11 [7-18] | 6 [3-11] |  |
| Plaque composition |  |  |  |  |
| Soft, n (%) | 317 (2) | 195 (3) | 122 (18) | 0.016* |
| Soft per patient | 1 [0-2] | 2 [0-4] | 1 [0-3] |  |
| Mixed, n (%) | 962 (7) | 555 (8) | 407 (6) | 0.004* |
| Mixed per patient | 4 [2-8] | 6 [3-9] | 3 [1-7] |  |
| Ruptured/thrombus, n (%) | 6 (<1) | 5 (<1) | 1 (<1) | 0.29 |
| Ruptured/thrombus, per patient | 0 [0-0] | 0 [0-0] | 0 [0-0] |  |
| Calcific, n (%) | 461 (3) | 351 (5) | 110 (2) | <0.001* |
| Calcific, per patient | 1 [0-4] | 3 [1-7] | 0 [0-1] |  |
| Non-calcified, n (%) | 1,285 (10) | 755 (12) | 530 (8) | <0.001* |
| Non-calcified, per patient | 7 [3-11] | 8 [4-13] | 6 [2-9] |  |
| Mann Whitney U test, Significance level *p* <0.05* |  |  |  |  |

Notably, amongst patients with SMuRFs and those who were SMuRF-less in the pooled ICM and NICM groups, there was no difference in total plaque burden, nor in the burden of non-calcified and calcified plaque (p>0.05 for all). **(Figure 6)**

**Figure 6:**
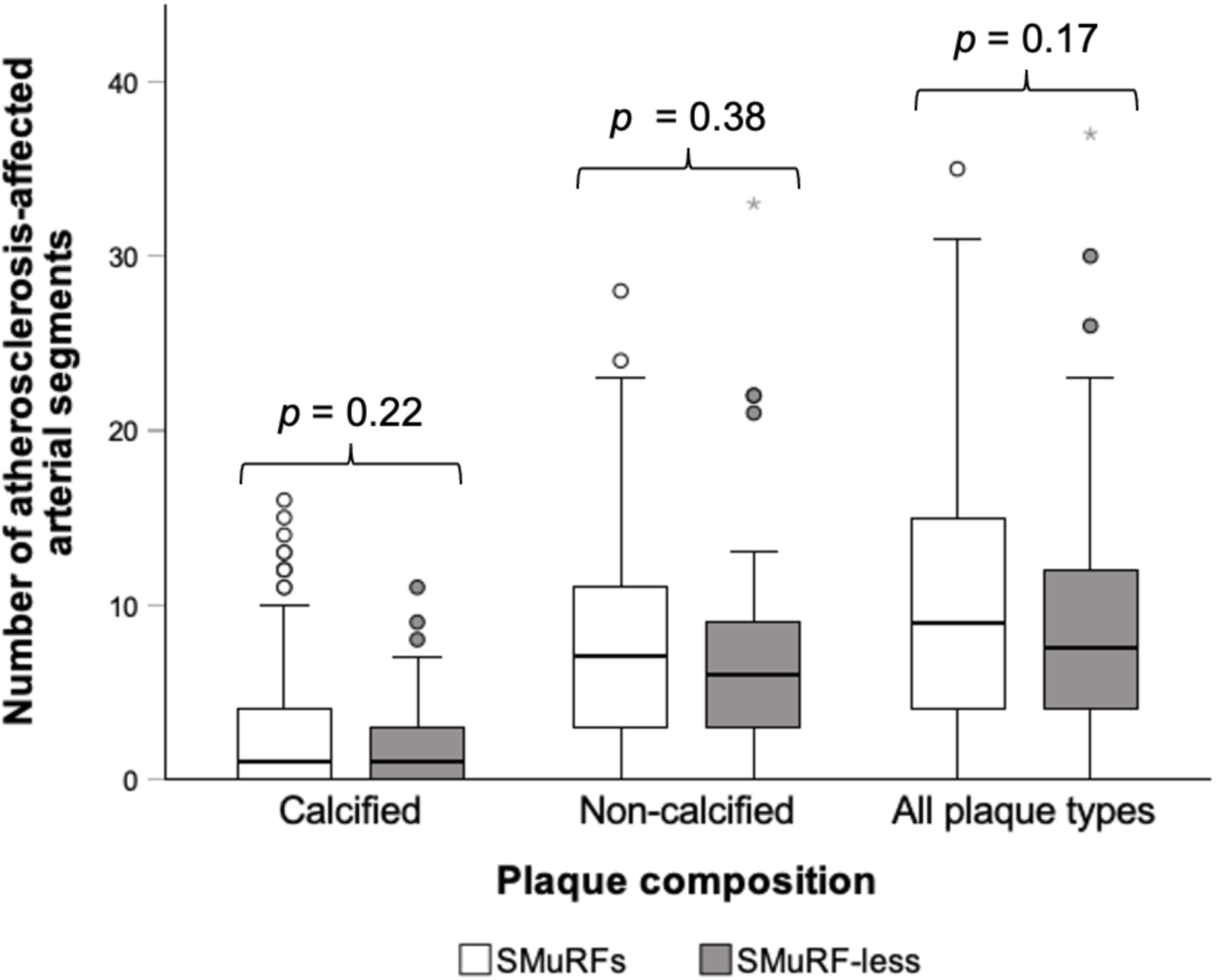
Non-calcified, calcified and total plaque burden in patients with SMuRFs and patients who were SMuRF-less.

Plaque burden was higher in ICM than in NICM. However, multivariable analysis incorporating sex, SMuRFs, residual cardiovascular risk factors and plaque-modifying pharmacological therapy showed that neither history of ICM nor history of at least one SMuRF remained associated with total, non-calcified or calcified plaque burden (p>0.05 for all). Total and non-calcified plaque burdens were not associated with atherosclerotic disease predictors. Notably, angiotensin receptor/neprilysin inhibitor use was inversely related to non-calcified plaque burden. Age and prior history of SMuRFs were positively associated with greater calcific plaque burden (age: *p* = 0.04; SMuRFs: *p* = 0.02), and baseline hsCRP was also independently associated with greater calcified plaque burden (p=0.002). LDL levels were inversely related to calcific plaque burden (*p* = 0.002). **(Table 5)**

**Table 5:** Multivariable linear regression.

| Predictor | $\beta$ | t | p |
| --- | --- | --- | --- |
| **Total plaque burden** |  |  |  |
| Male sex at birth | 0.12 | 0.71 | 0.48 |
| History of ICM | 0.11 | 0.65 | 0.52 |
| History of at least 1 SMuRF | 0.06 | 0.32 | 0.75 |
| Age (Years) | 0.23 | 1.33 | 0.2 |
| BMI (kg/m<sup>2</sup>) | 0.21 | 1.21 | 0.24 |
| Creatinine (μmol/L) | -0.07 | -0.46 | 0.65 |
| HbA1c (%) | -0.06 | -0.37 | 0.71 |
| Triglycerides (mmol/L) | 0.12 | 0.7 | 0.49 |
| LDL (mmol/L) | -0.32 | -1.84 | 0.08 |
| hsCRP (mg/L) | 0.12 | 0.7 | 0.49 |
| Cholesterol-lowering therapy§ | 0.05 | 0.3 | 0.77 |
| ARNI use | -0.36 | -1.97 | 0.06 |
| **Non-calcified plaque burden** |  |  |  |
| Male sex at birth | 0.03 | 0.17 | 0.87 |
| History of ICM | 0.2 | 1.07 | 0.29 |
| History of at least 1 SMuRF | -0.1 | -0.55 | 0.59 |
| Age (Years) | 0.13 | 0.71 | 0.48 |
| BMI (kg/m<sup>2</sup>) | 0.2 | 1.03 | 0.31 |
| Creatinine (μmol/L) | -0.07 | -0.44 | 0.67 |
| HbA1c (%) | 0.02 | 0.1 | 0.92 |
| Triglycerides (mmol/L) | 0.06 | 0.33 | 0.75 |
| LDL (mmol/L) | -0.16 | -0.85 | 0.4 |
| hsCRP (mg/L) | -0.08 | -0.46 | 0.65 |
| Cholesterol-lowering therapy§ | 0.001 | 0.01 | 0.99 |
| ARNI use | -0.41 | -2.1 | 0.044* |
| **Calcified plaque burden** |  |  |  |
| Male sex at birth | 0.25 | 1.85 | 0.07 |
| History of ICM | -0.14 | -0.99 | 0.33 |
| History of at least 1 SMuRF | 0.37 | 2.54 | 0.017* |
| Age (Years) | 0.31 | 2.17 | 0.038* |
| BMI (kg/m<sup>2</sup>) | 0.13 | 0.86 | 0.4 |
| Creatinine (μmol/L) | -0.06 | -0.43 | 0.67 |
| HbA1c (%) | -0.19 | -1.43 | 0.16 |
| Triglycerides (mmol/L) | 0.21 | 1.4 | 0.17 |
| LDL (mmol/L) | -0.5 | -3.44 | 0.002* |
| hsCRP (mg/L) | 0.46 | 3.37 | 0.002* |
| Cholesterol-lowering therapy§ | 0.13 | 0.92 | 0.36 |
| ARNI use | -0.04 | -0.29 | 0.78 |

Significance level $p < 0.05^*$
For categorical variables, 1 = Yes, 0 = No
§Cholesterol-lowering therapy included use of statins or ezetimibe, mono or dual therapy
ICM – Ischaemic cardiomyopathy SMuRF – Standard modifiable risk factor, includes: smoking, diabetes, hyperlipidaemia and hypertension BMI – Body mass index HbA1C – Glycated haemoglobin CVA – Cerebrovascular accident LDL – Low density lipoprotein hsCRP – High-sensitivity C-reactive protein ARNI – Angiotensin receptor-neprilysin inhibitor

## DISCUSSION

In patients with end-stage heart failure undergoing evaluation for heart transplantation, those with ICM exhibited greater extracardiac thoracoabdominal atherosclerotic plaque burden on pre-transplant CTA than patients with NICM. Although a higher atherosclerotic burden in ICM compared with NICM may seem expected, the magnitude of the difference in the presence of atherosclerosis, along with the absence of an association between plaque burden and SMuRFs between the groups, reveals a disconnection between conventional clinical risk assessment and the extent of systemic atherosclerosis. Hence, these findings suggest that thoracoabdominal plaque burden detected on routinely performed pre-transplant CTA is a meaningful marker of atherosclerotic disease severity beyond SMuRF-based assessment alone.

To the best of our knowledge, no studies have yet investigated differences in high-risk extracardiac plaque burden among patients with advanced heart failure from ischaemic and non-ischaemic causes. However, a substantial body of literature has established a definitive link between coronary and extracardiac, particularly in the middle-aged population. A prospective study from 1997, which enrolled 102 patients who underwent serial coronary angiography and ultrasound examination of several extracardiac arterial beds, including the thoracic aorta, carotid arteries, and femoral arteries, found that a high extracardiac atherosclerotic burden was positively associated with multivessel coronary disease, reflecting the systemic nature of atherosclerotic disease. (7)

The present current study found that a relatively high proportion of younger patients aged less than 55 years (male) or 65 years (female) with ICM and advanced heart failure had high-risk plaque in at least one extracardiac arterial segment, visible on thoracoabdominal CTA. Furthermore, in the presence of other standard and residual risk factors such as LDL, HbA1C and baseline hsCRP levels, we found that age was not associated with high-risk extracardiac plaque burden. Further prospective research is required to understand how the systemic effects of atherosclerotic disease differ between age groups.

### Implications for clinical management of patients with advanced heart failure and high-risk extracardiac atherosclerotic disease

The findings of this study contribute to the emerging discussion around standard modifiable risk factors as targets for atherosclerotic disease optimisation prior to heart transplantation in patients with advanced heart failure. This study found that no single modifiable or non-modifiable risk factor could be consistently associated with high-risk plaque burden. Additionally, the findings of this study challenge the conventional understanding of atherosclerosis as a disease associated most closely with aging. In our cohort of patients with end-stage heart failure, age was not a strong predictor of thoracoabdominal plaque burden. Notably, an overwhelming majority of patients with NICM (76/85, 89%) were found to have high-risk (soft, mixed or ruptured) plaque present in at least one visualised thoracoabdominal arterial segment.

Furthermore, in this cohort of patients with advanced heart failure, traditional SMuRFs showed little explanatory value for established atherosclerotic burden on CTA. Neither the presence of at least one SMuRF nor ischaemic versus non-ischaemic aetiology of heart failure independently associated with total or non-calcified plaque burden once age, renal function, lipids, inflammatory markers and pharmacological therapies were considered. SMuRF status also failed to relate to calcified plaque burden. Instead, systemic inflammation and ageing emerged as predictors of calcific atherosclerosis disease, with hsCRP and age showing associations with calcified plaque. Taken together, these data align with emerging reports of SMuRF-less patients experiencing acute coronary events despite an apparently low-risk profile (14). To this end, continued reliance on existing paradigms of risk stratification based on conventional risk factor assessment-alone could stand to further disadvantage the already vulnerable advanced heart failure population with potential impacts for their clinical outcomes.

### Thoracoabdominal CTA as a tool for determining risk of future adverse events

This study demonstrated that the evaluation and analysis of thoracoabdominal CTA imaging to interrogate extracardiac atherosclerotic plaque burden is a feasible method of identifying high-risk atherosclerotic plaque phenotypes which could signal the need for risk factor optimisation prior to transplant. Thoracoabdominal CTA offers many advantages over isolated imaging of the coronary or carotid arteries to determine clinical risk of future MACCE. Firstly, it is accessible in most centres and does not require ECG-gating or pre-medication, as would otherwise be required for CT coronary angiography. Secondly, in the advanced heart failure population, thoracoabdominal or whole-body CTA is performed routinely as part of pre-transplantation work-up to screen for malignancy. (15) Therefore, unanticipated insights from CTA assessment of extracardiac atherosclerotic plaque burden can reasonably be framed as an imaging biomarker of heightened clinical risk of MACCE while awaiting transplant, or of similarly systemic inflammatory disease processes which can occur following heart transplantation, such as recurrent CAD or cardiac allograft vasculopathy. (16, 17)

The integration of imaging markers of high-risk disease from distant arterial beds or even different tissue types, to complement or append focused imaging of atherosclerosis-prone arterial beds such as the coronary or carotid arteries may further refine risk stratification of atherosclerotic disease, beyond primordial risk factors. In a cohort study which included 5,059 postmenopausal women aged between 60-79 years, Iribarren and colleagues demonstrated that the presence of breast arterial calcification on screening mammography was independently associated with higher hazards of incident atherosclerotic disease and global cardiovascular events after adjustment for traditional risk factors. (18) It was found that women with BAC experienced consistently higher age-adjusted rates of myocardial infarction, stroke, cardiovascular death, and other major adverse cardiovascular and cerebrovascular events, indicating that this marker meaningfully enhances prediction of MACCE risk. (18) This finding supports the concept that atherosclerotic changes detected in one arterial territory can act as a surrogate marker of systemic disease burden and progression in other vascular beds. Consequently, incorporating such extra-coronary or extra-carotid markers into risk assessment may help identify individuals at heightened risk of future cardiovascular events who might otherwise be underestimated by conventional imaging confined to a single arterial bed.

A limitation of thoracoabdominal CTA is its relatively high radiation dose, which recent population models suggest could elevate the lifetime risk of neoplastic disease. Patients with orthotopic heart transplants are already at heightened cancer risk due to immunosuppressive therapy and may face additional risk from exposure to medical ionising radiation. (15, 19) CTA is also limited as a diagnostic tool by the inherent need for administration of intravenous iodinated contrast, so that pathologies affecting the vessel wall can be delineated from the vessel lumen. In this context, iodinated contrast administration is often contraindicated in the presence of renal dysfunction, which occurs frequently in subjects with advanced heart failure. (20)

Finally, CTA image quality and the ability to evaluate atherosclerosis-prone arterial regions is greatly reduced in the presence of implanted metallic devices, which cause metallic artefact and degrade spatial resolution in surrounding regions. Indeed, in this study, image quality in many examinations was reduced by the presence of implanted mechanical circulatory support devices (i.e. left ventricular assist devices) or cardiac resynchronisation therapy or pacemaker devices. Artefacts caused by implanted devices may be partially overcome by metal artefact reduction post-processing algorithms and the technical evolution of, though, given the high reliance on implanted devices in current end-stage HF heart failure management strategies before transplantation, metallic artefact from implanted cardiac devices could limit the translatability of thoracoabdominal CTA as a scalable screening tool in the advanced HF heart failure population. The emergence of next-generation photon-counting CT systems, with their improved spatial resolution and routine spectral information enabling high-keV virtual monoenergetic reconstructions, offer additional opportunities to further suppress metal artefact and blooming from cardiovascular implants in this setting. (21) Nevertheless, in the context of the current generation of CT scanners, the impacts of metallic artefact from implanted devices on image quality is a substantial limitation of thoracoabdominal CTA, which may disproportionately impact patients with advanced heart failure.

### Limitations

Although this study used a semi-qualitative design, it is acknowledged that quantitative methods for calculating plaque burden and characterising plaque from CTA data are well described. (22) The decision to use a semi-qualitative design in the present study stemmed from the inclusion of imaging data which was acquired over more than a decade using two CT scanners: from 2012 to 2020, the Aquilion ONE CT (Toshiba Medical Systems Corporation, now Canon Medical Systems Corporation, Otawara, Tochigi, Japan) and from 2020 to 2025 the Aquilion ONE Prism Edition CT (Toshiba Medical Systems Corporation, now Canon Medical Systems Corporation, Otawara, Tochigi, Japan). In this context, it was determined that a semi-qualitative method should be used to calculate and plaque burden and characterise plaque composition, as opposed to a strictly quantitative method which incorporated discrete measurements of plaque attenuation, which would have been prone to error due to innate differences in tube voltage or operation. (23, 24) Nevertheless, the semi-qualitative approach to plaque burden calculation may be considered an important limitation of this study. Additionally, there was a lack of coronary imaging data availability for all patients. As a result, this analysis, which centred on the evaluation of thoracoabdominal plaque burden, could not be complemented with corresponding CTA imaging data of coronary plaque. However, coronary angiography and echocardiography reports were used to classify patients into ICM or NICM groups and confirm prior history of OMI and revascularisation. Future studies could address this study’s limitations by incorporating a prospective design with longitudinal endpoints to evaluate the predictive value of thoracoabdominal plaque as a marker of increased clinical risk of MACCE, and a more comprehensive evaluation of cardiac, extracranial and thoracoabdominal arterial beds.

## CONCLUSIONS

These findings provide new insights into the systemic nature of atherosclerosis among patients with advanced heart failure awaiting transplantation. By applying thoracoabdominal CTA, we observed that atherosclerotic plaque burden extends well beyond the coronary or carotid vasculature and varies by cardiomyopathy type but not cardiovascular risk factor profile. By demonstrating that thoracoabdominal CTA is a feasible modality for evaluating extracardiac plaque burden, this study provides a valuable framework for identifying patients who require intensive pre- or post-transplant medical optimisation. Further research, which incorporates longitudinal endpoints and non-standard cardiovascular risk factors, may help establish the clinical significance of this finding in the context of the management of atherosclerotic disease in the advanced heart failure population.

## LIST OF ABBREVIATIONS

ASCVD: atherosclerotic cardiovascular disease
BMI: body mass index
CAD: coronary artery disease
CTA: computed tomography angiography
CTCA: computed tomography coronary angiography
hsCRP: high sensitivity C-reactive protein
HU: Hounsfield units
ICM: ischaemic cardiomyopathy
IHD: ischaemic heart disease
MACCE: major adverse cardiac and cerebrovascular events
NICM: non-ischaemic cardiomyopathy
OMI: obstruction myocardial infarction
SMuRFs: standard modifiable risk factors

## Data Availability

The data that support the findings of this study are available from the corresponding author upon reasonable request.

## Appendix A

Imaging protocol and scanning parameters for patients undergoing CTA for heart-transplant work-up.

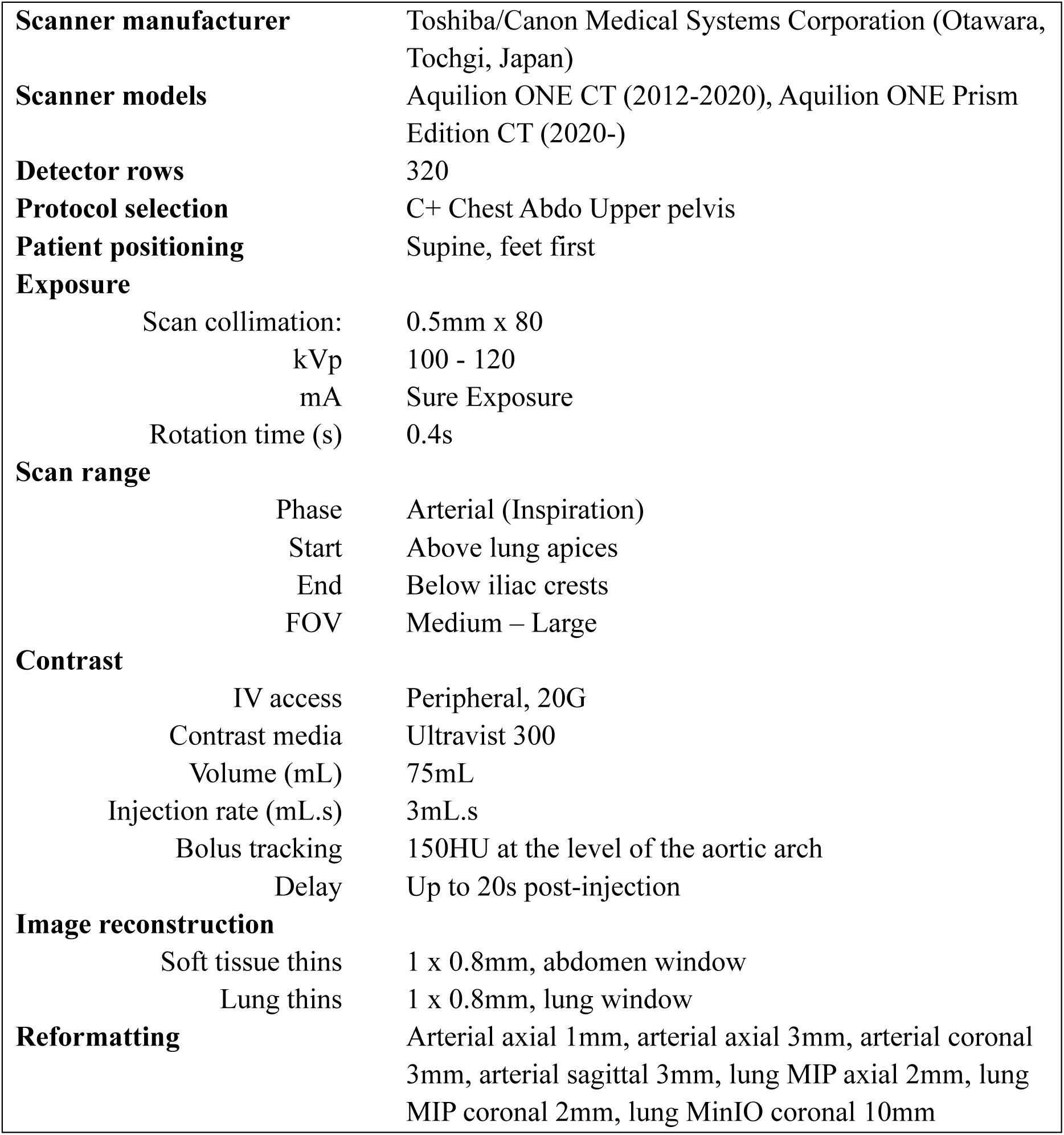

